# Yellow fever vaccination coverage among nomadic populations in Savannah region, Ghana; a cross-sectional study following an outbreak

**DOI:** 10.1101/2022.08.22.22279078

**Authors:** Abdul-Wahab Inusah, Gbeti Collins, Michael G Head, Peter Dzomeku, Shamsu-Deen Ziblim

**Author notes:** Correspondence author – Michael Head.

## Abstract

Yellow fever (YF) is a viral infection transmitted via mosquito bites. The disease is endemic in many African countries and Ghana has experienced frequent outbreaks. Vaccine coverage is often low in nomadic communities.

**Objective:** To evaluate YF vaccination coverage among nomadic population after the YF vaccination campaign in targeted communities in the Savanah region, Ghana.

**Study Design:** A community-based cross-sectional study, with a modified WHO vaccination coverage cluster survey was used to collect data from 2914 nomads in 414 nomadic households across 22 YF affected communities. Data were analyzed using Stata version 15. T-test analysis of variance was conducted to determine the statistical difference among different population groups.

**Results:** Out of the 2914 household members surveyed, 2342 (80%) were vaccinated against YF. There was a statistical difference between household size and household vaccination coverage with a mean difference of 1.38(p-value, <0.001). We found a statistical difference between YF vaccination coverage among the study population and that of the national coverage (88%) with a mean difference of 0.05(p-value =<0.001). About 94% of those vaccinated were able to show proof of vaccination with certified vaccination cards. The main reason for non-vaccination was household member/s travelling out of the district during the vaccination campaign.

**Conclusion:** YF vaccination coverage was below the national vaccination coverage, but within the WHO recommended threshold in obtaining herd immunity. The catch-up vaccination targeting hard-to-reach nomadic communities was necessary, in order to reduce likelihood of future outbreaks in these groups. Where resources allow, proactive monitoring of vaccine coverage and catch-up campaigns can help countries to meet 2026 international targets for YF elimination.

## Introduction

Yellow fever (YF) is a flavivirus-borne acute systemic illness spread by infected mosquitos of the Aedes and Haemogogus species (1). YF is difficult to diagnose since the symptoms and signs are similar to those of other diseases such as malaria, typhoid, dengue fever, and other haemorrhagic fever (2).

The disease can be transmitted to both human and non-human primates through Aedes mosquitos spp in Africa and *Haemagogus* spp. and *Sabethes* spp in Southern America (3). YF have three cycles of transmission; the Sylvatic cycle, which involved non-human primates (Aedes mosquitoes to monkey found in the forest). The intermediate cycle involves non-human primates, humans and *Aedes* spp. mosquitoes in African savannah settings, whilst the third cycle is the urban cycle involving primarily *Aedes aegypti* mosquitoes and humans in cities. However, the sylvatic and the intermediate cycle are the most common means of YF transmission in many high endemic African States (2).

There are international targets as part of the ‘Eliminate Yellow Fever Epidemics (EYE)’ initiative, with the primary outcome being to end yellow fever epidemics by 2026 (4). Annually, as of 2020, 200,000 cases of yellow fever are reported in Africa and South America, with 90% of these cases occurring in Africa resulting in an estimated 30,000 deaths (5). By region, West and Central Africa are reported to have the highest cases with about 300 probable and 88 laboratory-confirmed cases since 2021 (6), with frequent outbreaks being considered a local and global health threat (7). A majority of yellow fever cases in Africa are found among unvaccinated people living in the yellow fever endemic zones (8,9).

There is no known treatment or medication for yellow fever. however, vaccines have been a very effective tool in the prevention of yellow fever, with protection typically lasting for many years in over 80% of vaccinated individuals (10).

The estimated threshold for herd immunity and thus reduction in numbers and extent of the outbreak is thought to be around 80% (6). As of 2020, Africa’s YF vaccination coverage was 44% with the WHO citing this as one of the key challenges in controlling the incidence of yellow fever. (6). National coverage varies across West and Central Africa, for example, Ghana (88%), Congo (69%), Niger (67%), Cote D’Ivoire (69%), Cameroon (57%), Democratic Republic of Congo (DRC, 56%), Nigeria (54%), Central African Republic (CAR, 41%), and 35% in Chad (11). Overall, this often-low vaccination coverage makes the population susceptible to YF and the possible re-emergence of yellow fever in these countries (12,13).

Even though Ghana has a relatively high national yellow fever vaccination coverage (11), the country has seen a re-emergence of the disease. From October 15 to 27^th^ November 2021, 202 suspected cases of yellow fever including 70 confirmed positive cases with 35 deaths (17% case fatality ratio) were reported in four regions of Ghana (Upper West, Savannah, Bono, and Oti regions) (14). A vast majority of the positive cases reported were among the nomadic population suspected to be migrating from Nigeria into the Ghana Savanna forest reserves through its porous borders (14). The Ghana Health Service and partners since November 2021 have conducted focused YF vaccination campaigns in 80 targeted communities in the North and West Gonja Districts of the Savanna region. Efforts are also made to continue the catch-up vaccination campaigns using the routine immunization system. This study is part of the research efforts collating observations around knowledge, attitudes and practices towards yellow fever prevention and management in these communities (15). Here, the analyses cover YF vaccination coverage among nomadic groups in the West Gonja districts of the savannah of Ghana.

## Methods

### Study area, design and time period

A community-based cross-sectional survey was conducted between February to March 2022 among nomadic households in the 22 yellow fever outbreak communities in the West Gonja Municipal of the Savanna Region of Ghana. The Municipal has a population of 63,449 and is one of the seven districts of the Savanna region, and also serves as the administrative capital of the region(15). It is close to the borders of the Ivory Coast and Burkina Faso. The Municipal has a landmass of 4715.9sqkm, part of which is occupied by a protected forest reserve known as the Mole National Park(16).

### Study population, participants, and inclusion criteria

The study population was all members of nomadic households. Nomads in this study are members of a community without fixed habitation who regularly moves to and from for greener pasture for their livestock, farmland or both. Excluded populations included non-nomadic households and those ineligible for the YF vaccine (people less than 9 months, individuals allergic to egg products and pregnant women)(17). Household heads or spouses who were present in their households and gave informed consent, provided information for household members who were not present at the time of this study.

Using the yellow fever line list (a table containing detailed information on each case of a disease outbreak) obtained with permission from the Municipal Assembly Health Directorate, all communities with confirmed yellow fever cases were purposively selected for the study. In each community, community health volunteers(CHV) who previously participated in the most recent yellow vaccination campaign were used as focal persons to purposively identify all nomadic households for the research officers. Using the snowballing approach, consented household heads/spouses were also asked to identify other nomadic households for the research officers to approach.

### Sample size, Sampling Technique

A total population of 377 nomadic households were required to achieve the objective of the study at a 95% confidence level. This used the assumption of 50% vaccination coverage among the included communities, the margin of error of 5% and inflated the sample size by 5% for non-response and incomplete data entry.

### Data collection Tool, and Procedure

The study tool had two distinct sections; Section 1 obtained demographic information including gender, age (years), marital status, religious affiliation, household size, duration of stay, intention to relocate, and nationality. Section 2 collected data on households’ yellow fever vaccination coverage using a modified WHO post-vaccination cluster survey form(18). Evidence of vaccination was confirmed either via the vaccination card duly signed by a Ghana Health Service official, or a verbal report for missing cards, absent/dead household members.

The questionnaire was uploaded onto the Android smartphone App. (ODK) and pretested in the North Gonja District among similar study subjects. A face-to-face interview technique was used by trained research officers. This approach was adopted because it is thought that most of the study population is unable to read. The questions were read out in local dialects, predominantly Hausa, Fulani, Dagbani and Gonja. For each interview, an average of 25 minutes per household was used. there were 5 researchers officers carrying out the survey and data collection took 7 days to complete.

### Data analysis

The data collected was exported into Microsoft Excel 2019 for cleaning. The data analysis was carried out using the Stata version 15. Data were analysed using descriptive and inferential statistics.

After satisfying all assumptions (normality test) in running the t-test analysis, we conducted, a Pair sample T-test between Household size and Household Vaccination Coverage. One sample T-test between household vaccination coverage and National vaccination coverage. Finally, Independent Sample t-test between native and foreign nomadic yellow fever vaccination coverage. See S1 supplementary for an anonymized dataset. See S2 supplementary information for the full survey questions.

### Ethical consideration

Ethical approval for the study (UDS/RB/013/22) was obtained from the University for Development Studies (UDS) Research and Ethics Review Board (S3 supplementary). Also, permission was sought from the Savanna Regional Health Directorate through an introductory letter. The purpose of the study was explained to the study subjects. Respondents’ privacy and confidentiality of their responses were assured. Subjects were at liberty to discontinue the interview at any time. Respondents were required to provide their informed consent by way of signing or thumbprint before participating in the study. At the end of each interview, research officers spent five minutes educating the household on the signs and symptoms of yellow fever and various prevention strategies, including the benefits of vaccination.

## Results

### Socio-demographic characteristics of the study population

A total of 414 households participated in the survey. Among the study participants, 57.7% were males, with a mean age of 38.5± 13.1, and a range of 18 to 84 years. By relationship status, 91% of the participants were married, and the main occupation among participants was Herdsman (67.4%). By national status, 56% of the participants were foreign nomadic, and the majority migrated from the Benin republic. Table 1 describes the Socio-demographics of the participants.

**Table 1:**
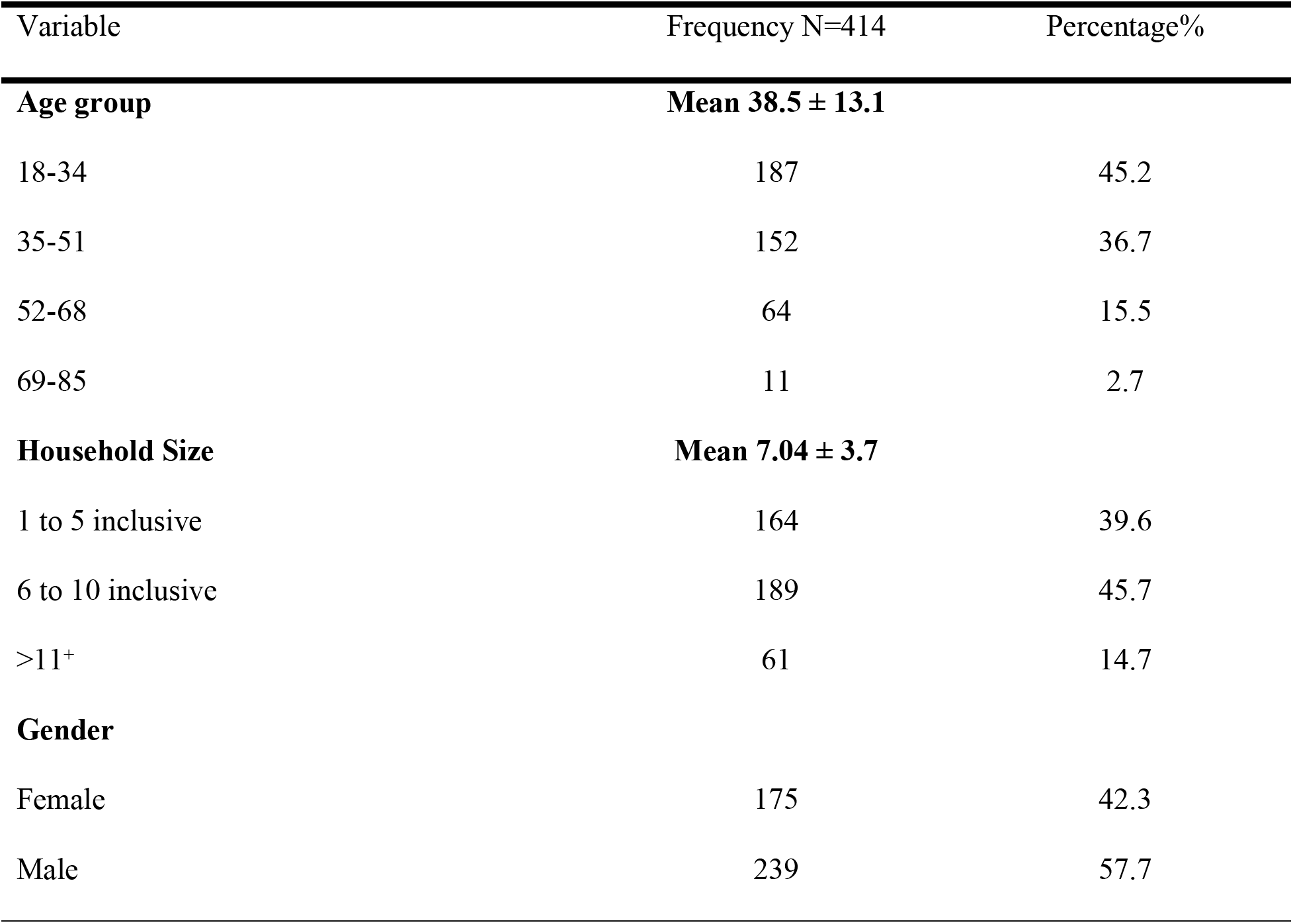
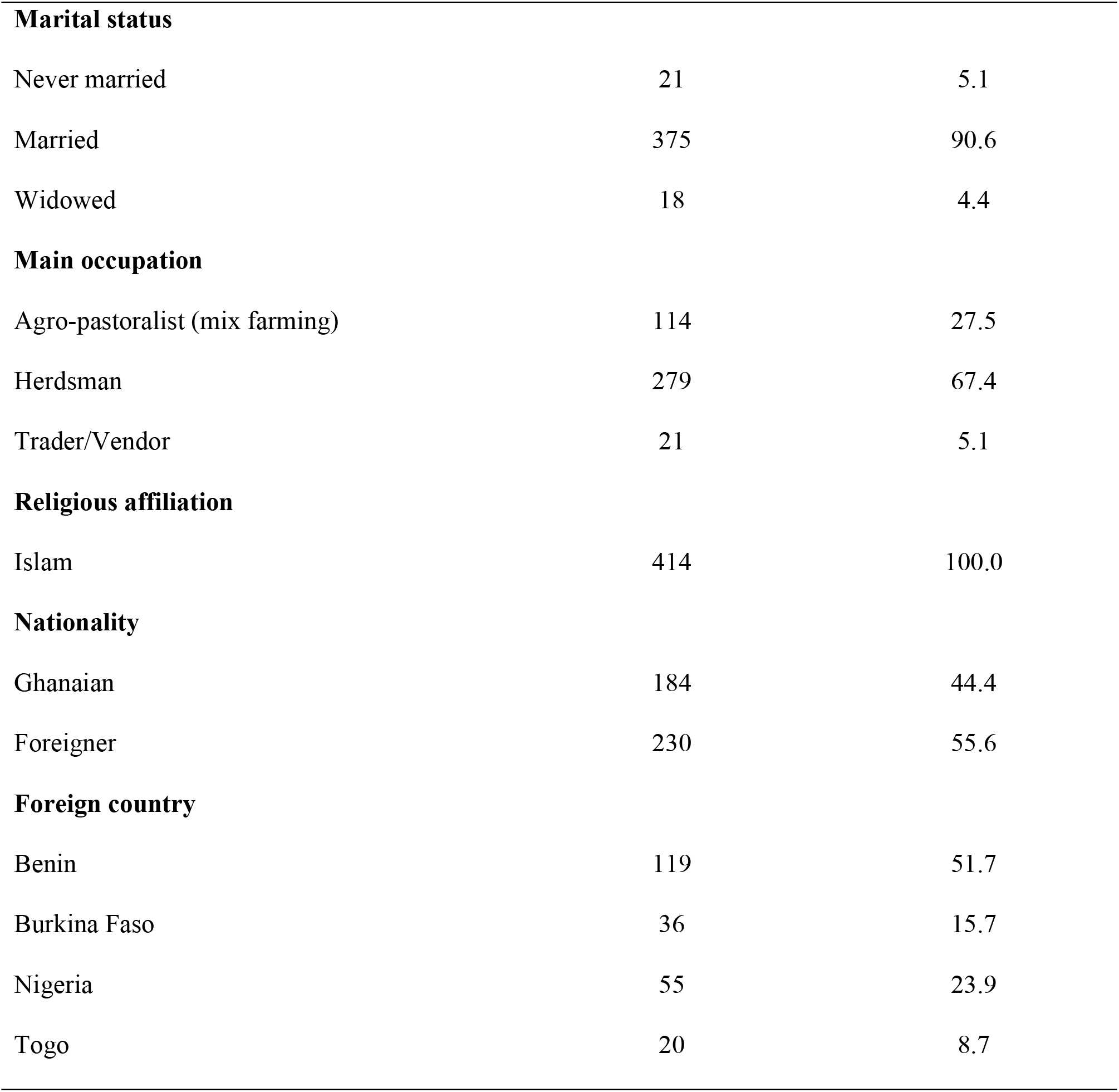
Sociodemographic characteristics of nomadic in the West Gonja Municipal Ghana

| Variable | Frequency N=414 | Percentage% |
| --- | --- | --- |
| **Age group** | **Mean $38.5 \pm 13.1$** |  |
| 18-34 | 187 | 45.2 |
| 35-51 | 152 | 36.7 |
| 52-68 | 64 | 15.5 |
| 69-85 | 11 | 2.7 |
| **Household Size** | **Mean $7.04 \pm 3.7$** |  |
| 1 to 5 inclusive | 164 | 39.6 |
| 6 to 10 inclusive | 189 | 45.7 |
| >11+ | 61 | 14.7 |
| **Gender** |  |  |
| Female | 175 | 42.3 |
| Male | 239 | 57.7 |
| **Marital status** |  |  |
| Never married | 21 | 5.1 |
| Married | 375 | 90.6 |
| Widowed | 18 | 4.4 |
| **Main occupation** |  |  |
| Agro-pastoralist (mix farming) | 114 | 27.5 |
| Herdsman | 279 | 67.4 |
| Trader/Vendor | 21 | 5.1 |
| **Religious affiliation** |  |  |
| Islam | 414 | 100.0 |
| **Nationality** |  |  |
| Ghanaian | 184 | 44.4 |
| Foreigner | 230 | 55.6 |
| **Foreign country** |  |  |
| Benin | 119 | 51.7 |
| Burkina Faso | 36 | 15.7 |
| Nigeria | 55 | 23.9 |
| Togo | 20 | 8.7 |

### Household heads Yellow fever Vaccination information

Table 2 shows participants’ information on yellow fever vaccination. Out of the 414 household heads interviewed, only 8(1.93%) said they personally have never received the yellow fever vaccination though other members of their households have received the vaccine. On proof of vaccination, 93.24% of the participants were able to show their vaccination cards with 4.82% using the verbal report as proof of vaccination as a result of missing cards. Participants were asked when they received their first yellow fever vaccination, significantly 399 participants (98.3%) received their yellow fever vaccination for the first time during the most recent vaccination campaign. Proportion of yellow fever vaccination was higher among foreign nomadic compared to the local nomadic(227/179). When considering perceptions around the vaccine, 67.5% indicated they believed the vaccine was effective or very effective; however, 25.1% suggested the vaccine was not effective at all.

**Table 2:** Study participants yellow fever vaccination information

| Variable | Frequency | Percentage |
| --- | --- | --- |
| **Have you ever been vaccinated against YF?** |  |  |
| No | 8 | 1.93 |
| Yes, the cards were not seen | 20 | 4.83 |
| Yes, the cards seen | 386 | 93.24 |
| **When did you receive your first YF vaccination?** |  |  |
| Before the most recent YF outbreak | 7 | 1.72 |
| During the just-ended YF vaccination campaign | 399 | 98.28 |
| **How effective is the vaccine in preventing yellow fever?** |  |  |
| Not effective at all | 102 | 25.12 |
| Slightly effective | 19 | 4.68 |
| Moderately effective | 11 | 2.71 |
| Very effective | 175 | 43.10 |
| Extremely effective | 99 | 24.38 |

### Household Members YF Vaccination Status

The total household members in the study were 2914, with 2342 household members ever receiving the yellow fever vaccine, this represents 80% vaccination coverage. Among those who have ever received the yellow fever vaccine, 2156 (92%) had their yellow fever vaccination cards showing evidence of vaccination. Those without the vaccination cards, either travelled with their cards or misplaced their cards at the time of this study.

### Reasons for non-vaccination against YF among household members

Several reasons were raised to explain why some household members (20%) failed to receive their vaccination (Figure 2). Closed to 62% (139) of the participants said those household members travelled out of town during the vaccination campaign period, 34% said lack of transportation to visit vaccination post(access), whilst 11% said fear of side effects after receiving the vaccine.

**Figure 1:**
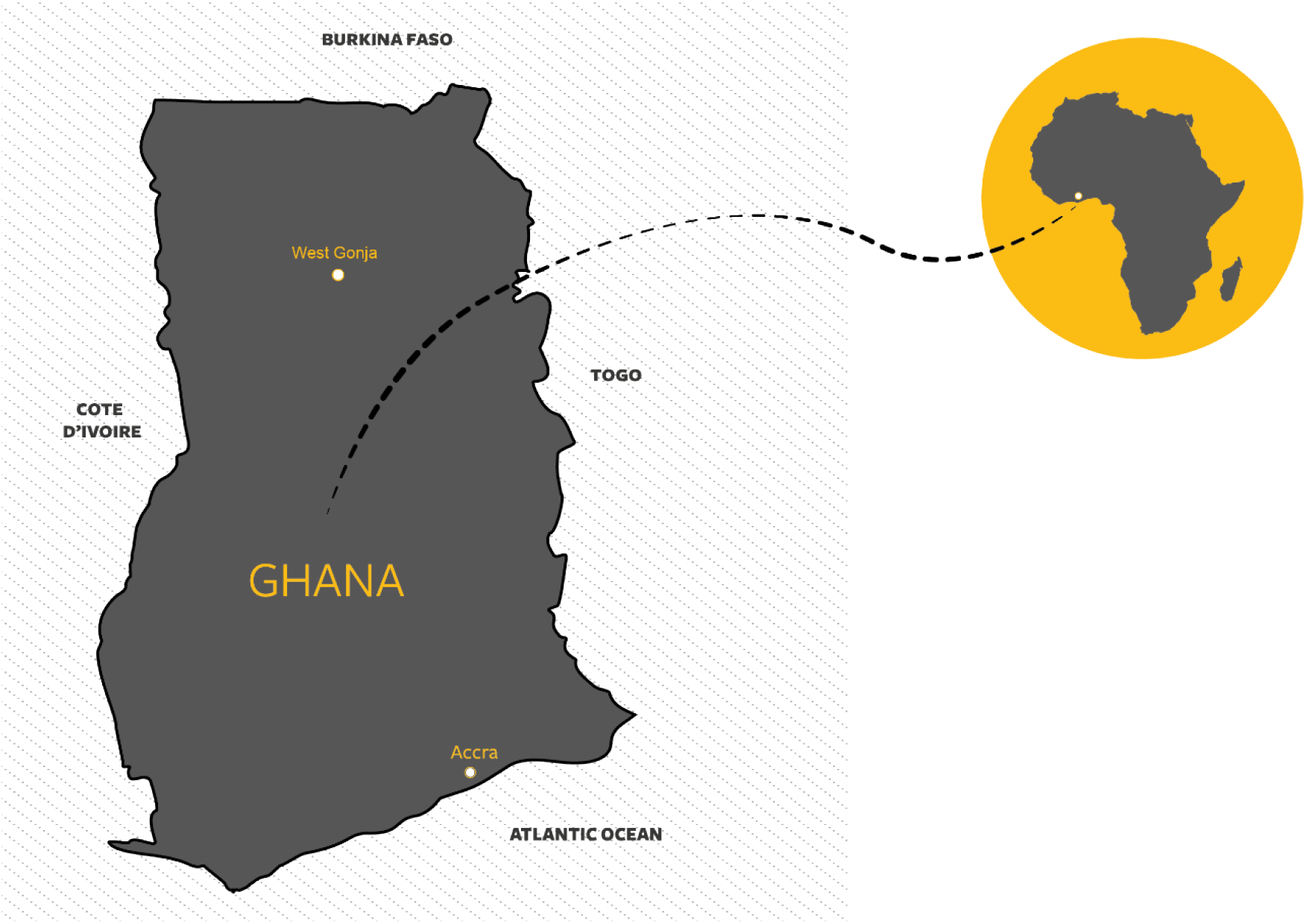
Map of Ghana showing the location of the West Gonja Municipal

**Figure 2:**
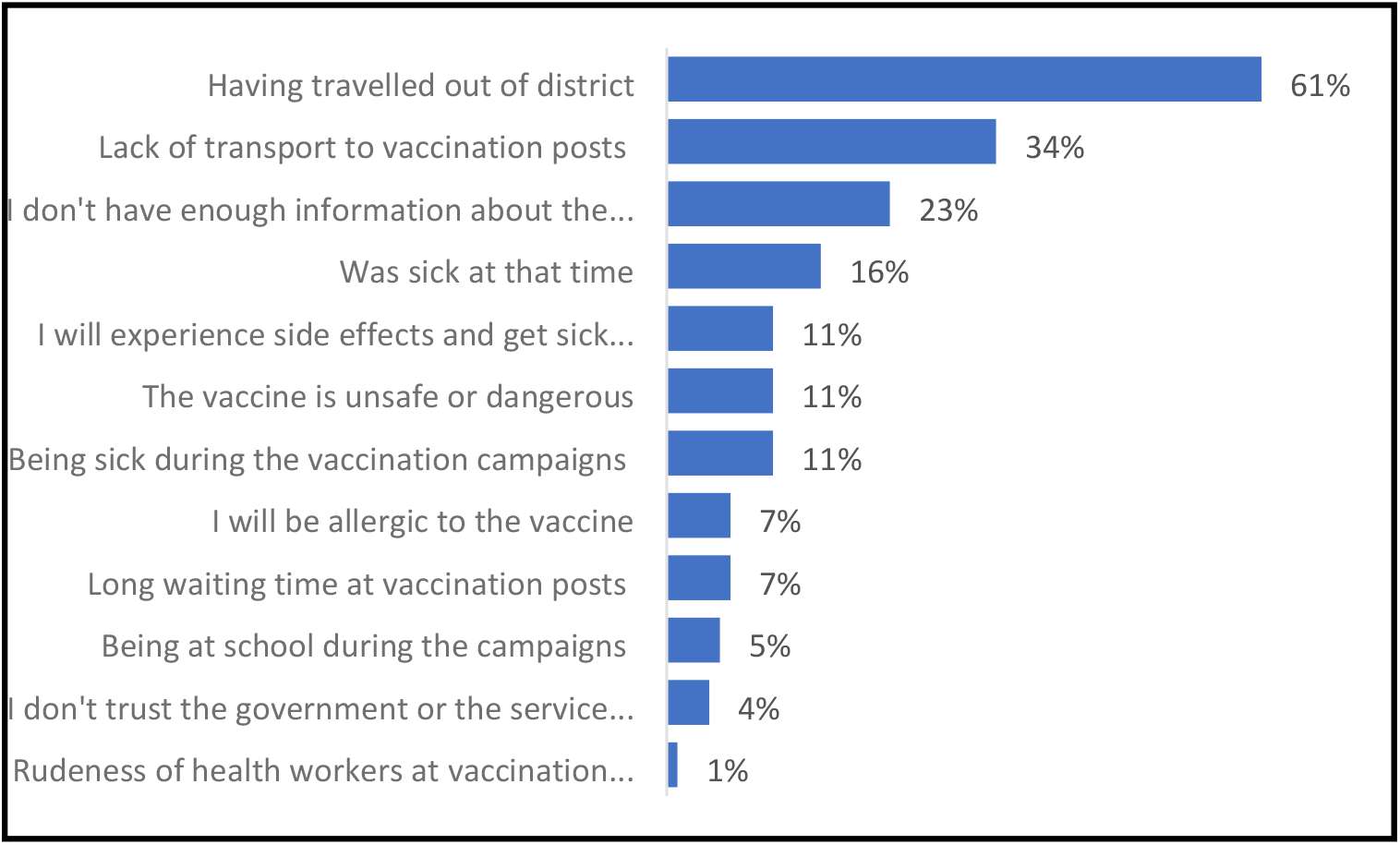
Reasons for yellow fever non-vaccination among study participants

### Analysis of yellow fever vaccination coverage in the study population Vs the National vaccination coverage (88%)

A one-sample t-test was conducted to compare the reported yellow fever vaccination coverage among the study population to the national yellow fever vaccination coverage which was 88% (14). The results showed a significant difference between the national yellow fever vaccination coverage and that of the study population. Vaccination coverage in the study population was (m= 0.83, SD=0.24). t(413)= −4.00, p-value =<0.001. The mean difference between the two groups was only 0.05. Compared to the national vaccination coverage, the result indicated a 5% (0.88-0.83) decrease in vaccination coverage in the study population as illustrated in Table 3.

**Table 3:** One sample T-test Analysis of Yellow Fever Vaccination Coverage between Study Population Vs National Vaccination Coverage

| Variable | Obs | Mean | Std. Err. | Std. Dev. | 95% Conf. Interval |
| --- | --- | --- | --- | --- | --- |
| Study pop. | 414 | 0.83 | 0.01 | 0.24 | 0.81 – 0.86 |
| Mean = mean (study pop.) |  |  |  |  | **t = -4.00** |
| Ho: mean(diff) = 0.88 |  | degrees of freedom = 414 |  |  |  |
| Ha: mean(diff) < 0.88 |  | Ha: mean(diff) != 0.88 |  | Ha: mean(diff) > 0.88 |  |
| Pr(T < t) = 0.00 |  | Pr(\|T\| > \|t\|) = **0.001** |  | Pr(T > t) = 1.00 |  |

### Analysis of Household Size Vs Household Yellow Fever Vaccination Coverage

Table 4 shows a pair sampled t-test analysis. This was conducted to evaluate the statistical difference between household size (2914), and household yellow fever vaccination coverage (2342). The results showed a significant difference between household size and household vaccination coverage. (m= 7.04, SD= 3.68) and against (m=5.66, SD= 3.13). t (413) = 11.83, p-value, <0.001. The mean decrease in the household population being vaccinated against yellow fever was 1.38 with a 95% confidence interval ranging from (1.15 to 1.61). The esta square statistics was (0.253) indicating a small effect size (19).

**Table 4:** Pair sample T-test Analysis of Household size Vs Household Vaccination Coverage

| Variable | Obs | Mean | Std. Err. | Std. Dev. | 95% Conf.<br>Interval |
| --- | --- | --- | --- | --- | --- |
| HHsize | 414 | 7.04 | 0.18 | 3.68 | 6.68 – 7.39 |
| HHvaccination | 414 | 5.66 | 0.15 | 3.13 | 5.36 – 5.96 |
| Diff | 414 | 1.38 | 0.12 | 2.34 | 1.15 – 1.61 |
| mean(diff) = mean (HHsize - HHvaccination) |  |  |  |  | **t = 11.83** |
| Ho: mean(diff) = 0 |  | degrees of freedom = 413 |  |  |  |
| Ha: mean(diff) < 0 |  | Ha: mean(diff) != 0 |  | Ha: mean(diff) > 0 |  |
| Pr(T < t) = 1.0000 |  | Pr(\|T\| > \|t\|) = **0.0000** |  | Pr(T > t) = 0.0000 |  |

### Analysis of Yellow Fever Vaccination Coverage between Native and Foreigner Nomadic

An independent sample t-test was conducted to compare yellow fever vaccine acceptance between Native nomadic and foreign nomadic As shown in Table 5, the results did not show any statistically significant difference in yellow fever vaccination coverage between the two groups. Native nomadic had (m= 5.44, SD=3.87) as against foreign nomadic (m= 5.087, SD= 2.43). t(183)= 1.023, p-value = 0.31. The mean difference between the two groups was only 0.35 with a 95% confidence interval of (−0.33 – 1.04).

**Table 5:** Independent Sample T-test analysis of Native nomadic Vs Foreign nomadic yellow fever vaccination coverage

| **Variable** | **Obs** | **Mean** | **Std. Err.** | **Std. Dev.** | **95% Conf.**<br>**Interval** |
| --- | --- | --- | --- | --- | --- |
| Native | 184 | 5.44 | 0.29 | 3.87 | 4.88 – 6.00 |
| Foreigner | 230 | 5.087 | 0.18 | 2.43 | 4.33 – 5.44 |
| Diff | 184 | 0.35 | 0.12 | 2.34 | -0.33 – 1.04 |
| mean(diff) = mean (HHsize - HHvaccination) |  |  |  |  | **t = 1.023** |
| Ho: mean(diff) = 0 |  |  | degrees of freedom = 183 |  |  |
| Ha: mean(diff) < 0 |  | Ha: mean(diff) != 0 |  | Ha: mean(diff) > 0 |  |
| Pr(T < t) = 0.85 |  | Pr(\|T\| > \|t\|) = **0.31** |  | Pr(T > t) = 0.15 |  |

## Discussion

This study was conducted in West Gonja municipal, after a focused vaccination campaign carried out over 80 communities of the West and North Gonja Districts of the Savanna Region. Our study found YF vaccination coverage to be 80% among the study population. Although this finding was below the national vaccination coverage of 88%(13), it was still within the WHO recommended threshold in attending herd immunity(13,18). We believed this vaccination coverage was a result of the compulsory nature of the vaccination among travellers (20) and the proactive approach towards community vaccination, as a result of the YF outbreak within the Savannah region. There were also additional opportunities for catch-up vaccinations outside of the immediacy of this yellow fever immunization campaign. Communities outside the vaccination campaign might have a lower coverage given the fact that the study population were scattered, highly mobile, and in the hinterlands.

As highlighted by Ghana’s national efforts, routine yellow fever vaccinations have the potential to reach much of the target population. Another example is in French Guiana where national vaccination coverage of 95%.

Proof of vaccination (here, a vaccination card or certificate) is often required to travel across borders in West Africa. In this study, card retention among those who received YF vaccination was 93.2%. This result was higher than in similar studies in Bolivia, Uganda and French Guiana (18,21,22). The higher vaccination card retention rate here was partly because of the short timeframe between immunization and study data collection, but perhaps also because this study population may be highly mobile and require proof of immunization whenever they need to move across borders(23).

Consistent with previous studies, the main reasons for not being vaccinated here included being out of the district during the vaccination campaign, lack of transport to the vaccination post and little information on the vaccination campaign(18,22). Access to healthcare is a common reason for vaccine hesitancy in Ghana and elsewhere, for example, in Malawi(24,25). This calls for consented efforts by health care workers to stay motivated in order to reach hard-to-reach communities to deliver quality health care service. Tailored health promotion to provide the right kind of information to their communities is key, along with logistics and planning of health services, ensuring they do not overlap or compete with each other.

The analysis did not show any statistically significant difference in vaccination coverage between the local nomads compared to international nomadic groups. This is probably due to the focused vaccination campaign adopting a house-house campaign strategy leading to every household having an equal chance of receiving the vaccine. The vaccination was free and taken into communities, with multiple chances to become vaccinated. This approach helps to reduce barriers to vaccine uptake, including cost, opportunity and access to healthcare(26).

WHO currently recommends a single dose of YF vaccine to provide lifelong immunity as a result, the vaccine has been introduced into the Expanded immunization programme for all children 9-12 months in high endemic countries like Ghana(26). Our study found a significant difference between household size and household members’ vaccination coverage. This indicates that not all household members were vaccinated during the vaccination campaign, and has implications for future transmission and outbreaks. Public Health authorities could consider catch-up vaccination campaigns in communities of low coverage in order to achieve the national YF vaccination threshold (14). Population and census data, and up-to-date information around nomadic household size, are important to prospectively inform planning and logistics around health services.

There are often inequities or inconsistencies in a vaccine rollout. Here, the study population still had a lower vaccine uptake compared to national coverage, and this was after a dedicated campaign to increase population protection. A greater difference was observed here than from a similar study in Bolivia which recorded a 4% difference when compared with national vaccination coverage (22). Studies like this one can support national-level data collection, by providing accurate estimates of vaccination coverage. National estimates can themselves be subject to error from inconsistencies or lack of high-quality data that include incorrect population estimates, incomplete tallying or reporting of vaccination doses (18,21,27).

### Limitation of the study

The fact that the interviews were conducted by proxy could introduce bias in the information provided. However, in all the interviews, household heads were recruited as the study participants who knew the demographic characteristic of their households and the household member vaccination status.

Again, recall bias may have occurred as a verbal report was one of the means of confirming vaccination status for those who could not produce their yellow fever vaccination cards. This was minimal as only 5% of household members could not produce their yellow fever vaccination cards. The evaluation was conducted immediately after the vaccination campaign reducing the tendency of recall bias.

## Conclusions

Our study found yellow fever vaccination in this nomadic population to be below the national threshold of 88%. Although the vaccination coverage was within the WHO recommended threshold(80%) to eliminate human-to-human transmission of the disease(21), catch-up and revaccination activities were necessary targeting those hard-to-reach areas to attain the national threshold to contain a possible transmission of the disease among the unvaccinated population. These study findings show that nomadic populations may be unvaccinated against yellow fever and thus regular catch-up campaigns should target these demographics. Their mobility across endemic areas makes them vulnerable to infection. Future research should consider conducting a nationally representative survey to determine the yellow fever vaccination coverage in nomadic populations, as well as considering similar approaches to assessing effective health promotion content that increases their access to healthcare.

## Data Availability

The anonymised dataset is available in the supplementary information alongside this manuscript.

## Competing interests

The authors declare that they have no competing interests

## Acknowledgements

We acknowledge the Regional Health Directorate, Community leaders, Community Health Volunteers, data collectors and individuals who participated in the survey. We are grateful to Alhaj Mohammed Tahidu of the Northern Regional Health Directorate for supporting us in obtaining the line list used for this study.

## Author Contributions

Conceptualization: AWI

Data curation: AWI, MH.

Formal analysis: AWI, GC, SDZ, MH.

Investigation: AWI.

Methodology: AWI, GC, SDZ, PJK.

Project administration: AWI Supervision: AWI

Writing – original draft: AWI, GC

Writing – review & editing: AWI, GC, MH, SDZ, PJK,

All authors approved the submitted and final version of this manuscript.

## Notes

### Competing Interest Statement

The authors have declared no competing interest.

### Funding Statement

The authors received no specific funding for this work.

### Author Declarations

Ethical approval for the study (UDS/RB/013/22) was obtained from the University for Development Studies (UDS) Research and Ethics Review Board.

